# Randomized Controlled Trials of Early Ambulatory Hydroxychloroquine in the Prevention of COVID-19 Infection, Hospitalization, and Death: Meta-Analysis

**DOI:** 10.1101/2020.09.30.20204693

**Authors:** Joseph A. Ladapo, John E. McKinnon, Peter A. McCullough, Harvey A. Risch

## Abstract

**Objective:** To determine if hydroxychloroquine (HCQ) reduces the incidence of new illness, hospitalization or death among outpatients at risk for or infected with SARS-CoV-2 (COVID-19).

**Design:** Systematic review and meta-analysis of randomized clinical trials.

**Data sources:** Search of MEDLINE, EMBASE, PubMed, medRxiv, PROSPERO, and the Cochrane Central Register of Controlled Trials. Also review of reference lists from recent meta-analyses.

**Study selection:** Randomized clinical trials in which participants were treated with HCQ or placebo/standard-of-care for pre-exposure prophylaxis, post-exposure prophylaxis, or outpatient therapy for COVID-19.

**Methods:** Two investigators independently extracted data on trial design and outcomes. Medication side effects and adverse reactions were also assessed. The primary outcome was COVID-19 hospitalization or death. When unavailable, new COVID-19 infection was used. We calculated random effects meta-analysis according to the method of DerSimonian and Laird. Heterogeneity between the studies was evaluated by calculation of Cochran Q and I^2^ parameters. An Egger funnel plot was drawn to investigate publication bias. We also calculated the fixed effects meta-analysis summary of the five studies. All calculations were done in Excel, and results were considered to be statistically significant at a two-sided threshold of P=.05.

**Results:** Five randomized controlled clinical trials enrolling 5,577 patients were included. HCQ was associated with a 24% reduction in COVID-19 infection, hospitalization or death, P=.025 (RR, 0.76 [95% CI, 0.59 to 0.97]). No serious adverse cardiac events were reported. The most common side effects were gastrointestinal.

**Conclusion:** Hydroxychloroquine use in outpatients reduces the incidence of the composite outcome of COVID-19 infection, hospitalization, and death. Serious adverse events were not reported and cardiac arrhythmia was rare.

**Systematic review registration:** This review was not registered.

## Introduction

The coronavirus disease 2019 (COVID-19) pandemic, caused by the severe acute respiratory syndrome coronavirus 2 (SARS-CoV-2), has killed more than 200,000 people in the U.S. and 970,000 people worldwide as of mid-September 2020. Clinical studies testing the effectiveness of therapies for COVID-19 have primarily focused on hospitalized patients late in the course of illness, with evidence from randomized trials thus far favoring low-dose daily dexamethasone^1^ and possibly remdesivir.^2^ Rigorous randomized trials of therapy for COVID-19 in outpatients are limited. However, early treatment holds considerable value, considering the benefits of preventing disease progression and hospitalization, and the potential for accessible outpatient therapy to ameliorate the extraordinary social and economic burden associated with the pandemic.

Hydroxychloroquine (HCQ), as an antimalarial/anti-inflammatory drug, may be especially pertinent for treating COVID-19 infection in early stages of the disease. HCQ impairs endosomal transfer of virions within human cells.^3^ It is also a zinc ionophore, conveying zinc intracellularly to block the SARS-CoV-2 RNA-dependent RNA polymerase, which is central to the virus’s ability to replicate. A large number of non-randomized but controlled trials have now shown benefit of HCQ when used early for treatment of high-risk outpatients.^4 5 6 7 8 9^ Various randomized trials of HCQ for pre-exposure prophylaxis, post-exposure prophylaxis, and outpatient treatment have been performed. Individually, these clinical trials have yielded estimates of effectiveness that have not reached statistical significance.

One factor that may have contributed to the absence of statistical significance is early study termination. While the trials performed in Spain came close to accruing their intended number of patients (2,850 of 3,040 in total), the trials performed in Minnesota were administratively stopped prematurely by the investigators, before less than half of their enrollment goals had been met (1,312 of 3,000 in trial NCT04308668 and 1,496 of 3,500 in trial NCT04328467). In addition, all outpatient trials published so far have primarily enrolled healthy adults at low-risk of developing severe COVID-19 illness or serious outcomes. These considerations limit the statistical power of any individual outpatient randomized HCQ trial to yield precise estimates of efficacy. Moreover, these factors may explain why the individual trials have not rejected the null hypothesis. However, given their limitations, the appropriate conclusions would have been that the results observed, in the beneficial direction, just did not reach customary statistical significance.

The objective of our study was to determine, in randomized controlled trials, whether hydroxychloroquine use reduces hospitalization and mortality risks among outpatients with COVID-19. In otherwise eligible clinical trials for which these outcomes were unreported or uninformative, we considered development of COVID-19 illness to be the relevant clinical outcome for demonstrating drug benefit, as it was the main goal of these particular studies. We systematically searched the scientific literature and performed a meta-analysis of only randomized trials.

## Methods

We report this systematic review following the guidelines of the Preferred Reporting Items for Systematic Reviews and Meta-Analyses (PRISMA) checklist. We searched 5 medical bibliographic databases for relevant RCTs of outpatient HCQ use, with search terminology, ((hydroxychloroquine[Title]) AND (covid[Title] OR covid-19[Title] OR coronavirus[Title] OR SARS-CoV-2[Title] OR 2019-nCoV[Title])) AND (randomized[Text Word]). We initially identified 90 results in PubMed, 13 results in medRxiv, 77 results in Medline/Embase, and 72 results in the Cochrane database. In addition, we searched the PROSPERO database with search terminology hydroxychloroquine AND (Intervention OR Prevention) and identified 73 results. All of these papers were manually searched to identify RCTs of HCQ use in outpatients. We also examined reference lists in recent large meta-analysis papers. The database search identified 5 relevant studies as shown in Table 1. No additional studies were found in reference lists.

**Table 1.** Randomized Controlled Trials of Hydroxychloroquine Use in COVID-19 Outpatients

| First Author | Register ID | Treatment comparison<br>(Exposed n/<br>Control n) | Treatment schedule | Outcomes<br>(Exposed n/<br>Control n) | Age<br>(median<br>or mean) | Location | Blinding |
| --- | --- | --- | --- | --- | --- | --- | --- |
| Boulware DR<sup>14</sup> | NCT04308668 | 414/407 | 800mg once, then 600 mg 6 to 8 hours later, then 600mg qd for 4 more days | New COVID-19 infection<br>49/58 | 40 | US, Canada | No. Placebo was folic acid, recognizable |
| Skipper CP<sup>12</sup> | NCT04308668 | 212/211 | 800mg once, then 600 mg 6 to 8 hours later, then 600mg qd for 4 more days | Hospitalization or death<br>5/10 | 40 | US, Canada | No. Placebo was folic acid or lactose |
| Rajasingham R<sup>13</sup> | NCT04328467 | 989/494 | 400mg bid day-1, then 400mg once or twice weekly for 12 weeks | New COVID-19 infection<br>0.73 (0.48 to 1.09)* | 41 | US, Canada | No. Placebo was folic acid, recognizable |
| Mitjà O<sup>11</sup> | NCT04304053 | 169/184 | 800mg day-1, then 400 mg qd for 6 days | Hospitalization<br>8/12 | 42 | Catalonia, Spain | No. No placebo used |
| Mitjà O<sup>10</sup> | NCT04304053 | 1,197/1,300 | 800mg day-1, then 400 mg qd for 6 days | Death<br>5/8 | 49 | Catalonia, Spain | No. No placebo used |

\* For the Rajasingham<sup>13</sup> study, the new infection hazard ratio and its 95% confidence limits from Cox regression of its two HCQ groups combined vs placebo was given in the study, as shown.

Because these RCT studies were carried out in generally low-risk individuals, they were designed for outcomes of moderate clinical interest but not for serious disease consequences, which are few in such individuals. In addition, the studies were also terminated early by their investigators, lowering statistical power even further. The most important clinical outcome is mortality, and for outpatients, hospitalization conveys high risk of mortality. Thus, where studies observed more than 1 unexposed deceased or hospitalized subject, we used mortality or hospitalization or the two together as the outcome of interest for our meta-analysis.^10 11 12^ In studies where this was not the case, we used the study principal outcome (newly occurring COVID-19 infection, which is in the causal pathway to COVID-19 hospitalization) as defined by the study investigators.^13 14^

For analysis, from each study report we extracted the numbers of hospitalizations or deaths or newly occurring COVID-19 infections, as appropriate, for subjects given HCQ and for control subjects (Table 1). In one of the studies,^13^ the authors provided the new infection hazard ratio and its 95% confidence limits from Cox regression of its two HCQ groups combined vs placebo, and we used those values for our analysis. The other studies did not provide Cox regression results, thus we calculated relative risks and their 95% confidence limits from the numbers of subjects. In addition, we assessed medication side effects and adverse reactions.

We calculated random effects meta-analysis summaries of the five studies, with between-studies variance component according to the method of DerSimonian and Laird.^15 16^ We evaluated heterogeneity between the studies by calculation of Cochran Q and I^2^ parameters. An Egger funnel plot was drawn for the studies which suggested slight asymmetry of the smallest studies (data not shown). We tested this diagnostically by adding a hypothetical opposite study of the smallest included study to our meta-analysis, but it did not change the results at 2 digits of precision. We also calculated the fixed effects meta-analysis summary of the five studies. All calculations were done in Excel, and we considered results to be statistically significant at a two-sided threshold of P=.05.

## Results

The meta-analysis result for the five studies is shown in Figure 1. The heterogeneity of these studies was zero (P=.92), thus the fixed-effects and random-effects calculations provide congruent summary results as shown in the figure, 24% reduced outcome risk for the composite of COVID-19 infection, hospitalization, and death, P=.025.

**Figure 1.**
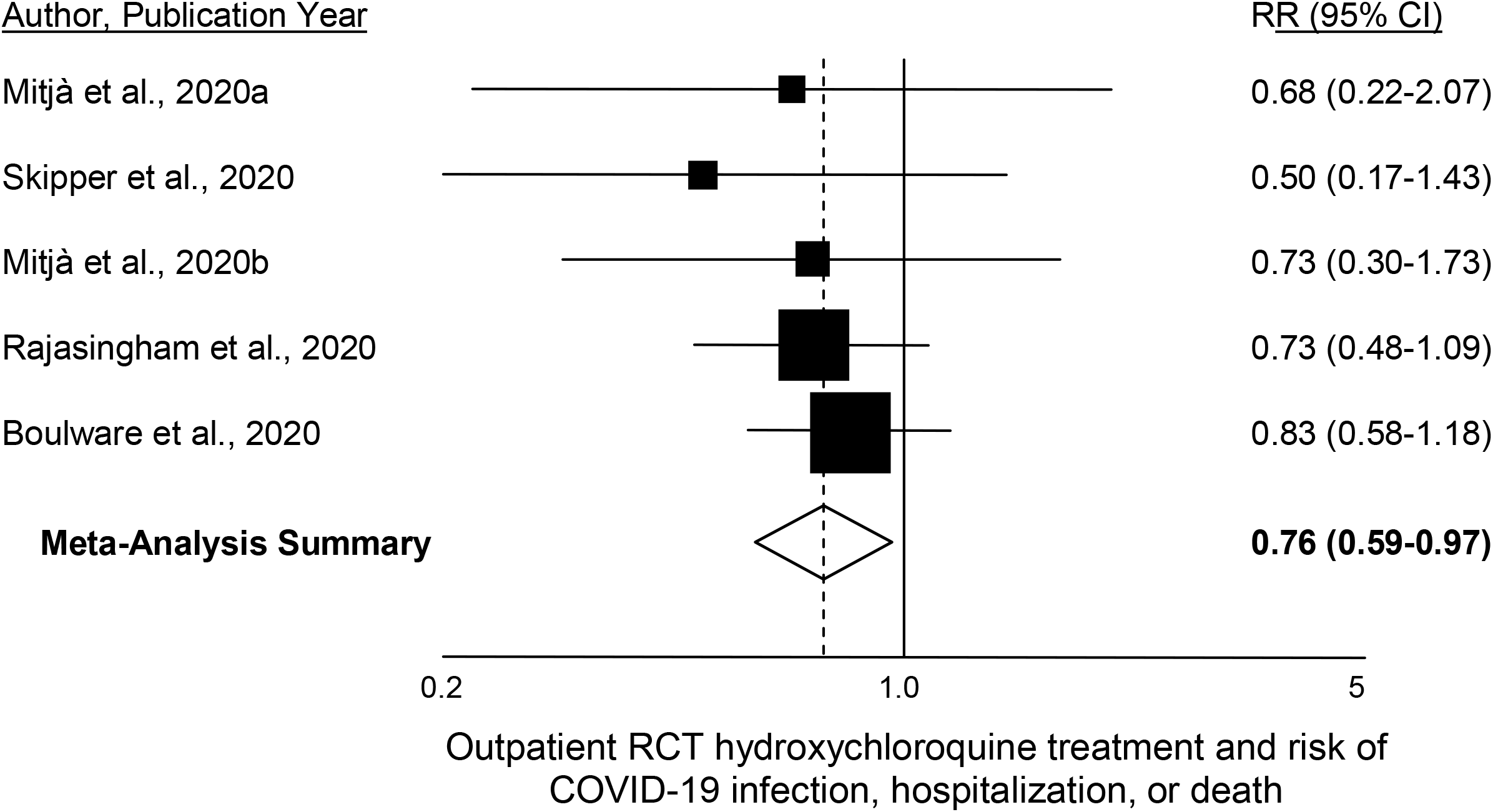
Meta-analysis funnel plot of 5 RCTs of outpatient hydroxychloroquine use. Area of the black squares is proportional to the study weight in the meta-analysis. The between-study heterogeneity is zero, thus the summary result is identical for fixed-effects and random-effects calculations. The study outcomes considered in the figure are, Mitjà et al., 2020,^10^ death; Mitjà et al., 2020,^11^ hospitalization; Skipper et al., 2020,^12^ hospitalization or death; Rajasingham et al., 2020,^13^ new COVID-19 infection; Boulware et al., 2020,^14^ new COVID-19 infection.

Watanabe^17^ observed that antiviral medication use starting many days after acquisition of infection should not be useful for prevention, and reanalyzed the Boulware et al. study according to specific HCQ start day after infection exposure. For medication use starting within 2 days after infection exposure, his analysis yields a relative risk of new COVID-19 infection of 0.64 (95%CI 0.36 to 1.14). Using this value in the meta-analysis instead of the Boulware et al. grouping of days 1-4 as included in Figure 1, gives a summary relative risk favoring hydroxychloroquine of 0.68 (95%CI 0.51 to 0.91), P=.0097.

As we have noted (Table 1), all of the studies involved young- to middle-aged adults who are generally at low-risk of COVID-19 progression and mortality. However, in the Spanish cluster-randomization study,^10^ 293 nursing-home residents at high-risk were also included in the patient mix. In these individuals, HCQ use for post-exposure prophylaxis reduced the risk of developing PCR-confirmed COVID-19, by half: relative risk 0.49 (95%CI 0.21 to 1.17).

In our assessment of side effects and adverse events (Table 2), four studies assessed cardiac arrhythmia, which rarely occurred. Specifically, three of the four studies reported no cardiac arrhythmias and the fourth study reported a cardiac arrhythmia in 1 out of 936 patients receiving HCQ versus 1 out of 469 patients in the control group. QT prolongation was not reported by any study. There were no serious cardiac adverse events and no trial was stopped early due to safety concerns. The most common side effects reported across all studies were gastrointestinal.

**Table 2.**
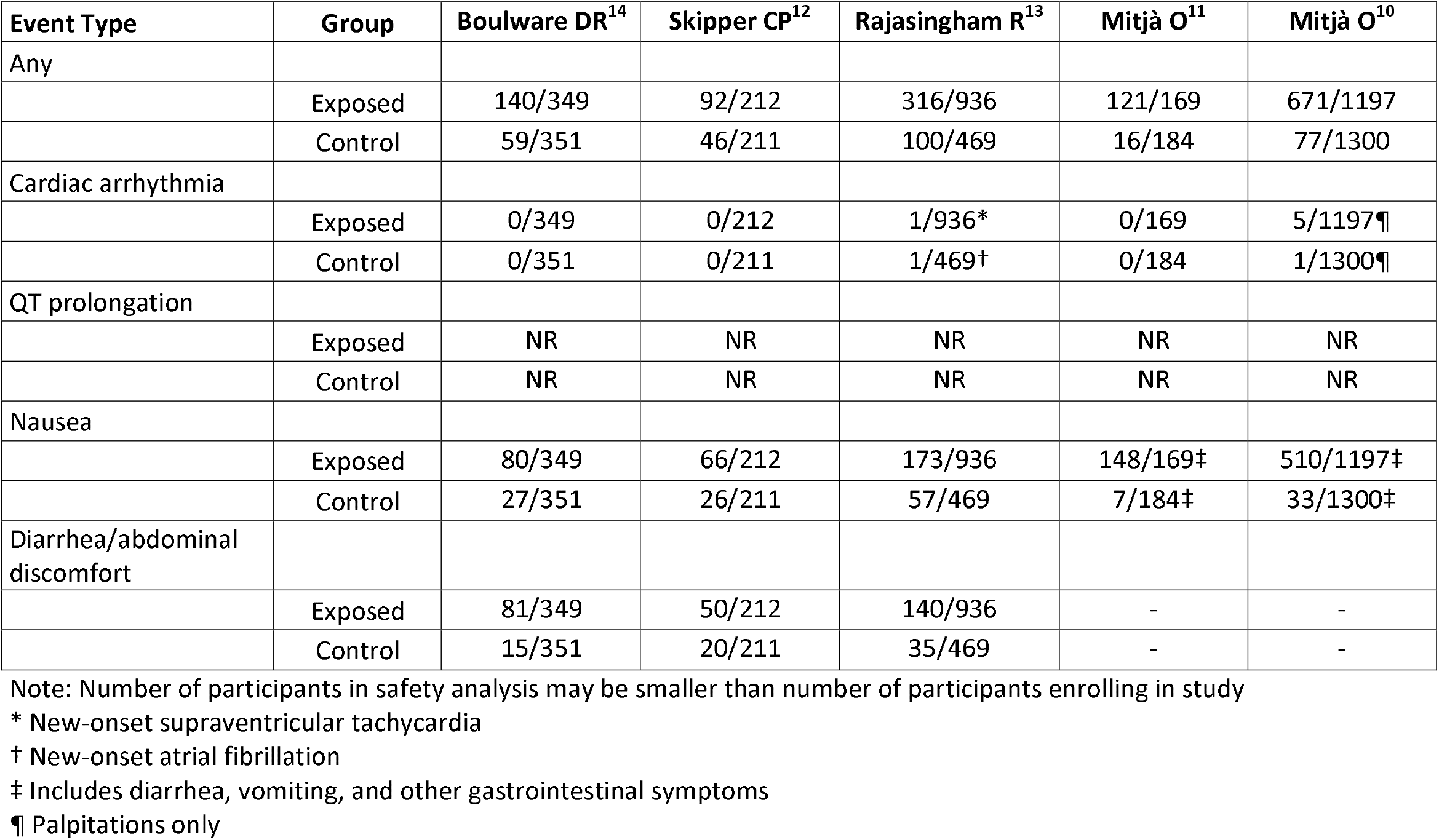
Side Effects and Adverse Events in Randomized Controlled Trials of Hydroxychloroquine Use in COVID-19 Outpatients

## Discussion

In this meta-analysis of 5 randomized clinical trials including 5,577 patients in the United States, Canada, and Spain, we found that outpatient use of HCQ for prophylaxis or early treatment of COVID-19 significantly reduced the composite of infection, hospitalization, and death. The magnitude of the benefit varied across studies, but was consistently present in each clinical trial, with varying levels of statistical precision. Our meta-analysis finding of a benefit from early HCQ prophylaxis or early treatment was significant despite somewhat diverse study characteristics that reduced the likelihood of detecting an effect, such as early study termination (in 3 studies) or testing shortages that forced reliance on suspected diagnoses. Our meta-analysis will be updated as the results of additional randomized trials of outpatient HCQ use are published.

A central hypothesis in the evaluation of outpatient therapies for COVID-19 is that anti-viral treatment is more effective the earlier by day in the disease course it is started. Among 1,840 clinical trials for COVID-19, it was recently reported that 62% involved patients who are hospitalized.^18^ These patients tend to be in late stages of the COVID-19 disease course, and their pathophysiology differs from the pathophysiology of patients in earlier stages of illness, or in a pre-exposure or post-exposure prophylaxis stage. Early disease is flu-like, versus hospitalized disease that is generally characterized by pneumonia or acute respiratory distress syndrome (ARDS). Patients are not hospitalized for flu-like symptoms. Although some randomized trials of HCQ have yielded unfavorable findings for hospitalized patients, these findings cannot be translated to the outpatient setting. Our meta-analysis shows that HCQ’s effectiveness is most evident in early ambulatory patients with COVID-19.

Our findings are consistent with the results of several non-randomized controlled evaluations of HCQ outpatient therapy for COVID-19, which have reported substantial effectiveness.^4 5 6 7 8 9^ In addition, a number of studies examining HCQ use in the hospitalized setting—particularly use starting within 24-48 hours of admission—have also shown benefit.^19 20 21 22 23 24^ That modern, well-conducted non-randomized trials and observational studies would yield results similar to those of RCTs is not surprising as it is the common finding across numerous medical disciplines.^25 26^

A review of the safety data reported by each trial indicates that cardiac arrhythmias were rare. In addition, no serious cardiac side effects were reported. This is similar to the lack of such events in the thousands of patients in all of the non-randomized controlled outpatient HCQ trials cited above. Appropriate off-label use of HCQ is nevertheless a clinical decision that incorporates considerations of individual contraindications, predispositions, correctable electrolyte abnormalities and possible ECG or laboratory testing in certain cases.

The potential role of early outpatient treatment for COVID-19 with HCQ and other agents was recently described.^27^ While our meta-analysis demonstrates that hydroxychloroquine reduces adverse clinical outcomes among patients with or at risk for COVID-19, alternative outpatient therapies may be effective and would benefit from further investigation and meta-analysis. These include zinc, prednisone, colchicine, and HCQ combination therapy with azithromycin, doxycycline, or favipiravir.

Our meta-analysis has limitations common to evaluations of preliminary trials that themselves have shortcomings, including truncated sample sizes and lack of placebo control. The clinical trials used different measures to define the primary outcome. To address this limitation, we prioritized assessing COVID-19 death and hospitalization, which are meaningful clinical outcomes, and used the authors’ listed primary outcome when death or hospitalization were unavailable. Whether the degree of benefit for risk of developing COVID-19 illness compared to the risk of mortality or hospitalization are comparable in magnitude is a potential but minor limitation, in that combining them in meta-analysis averages the benefit over these various types of patient outcomes. If the meta-analysis demonstrated appreciable heterogeneity between these types of studies, their outcome differences might be an explanation. If the meta-analysis demonstrated a null or statistically insignificant result, it could be argued that the outcome differences counteracted each other. Neither of these circumstances occurred, thus the combination of risk reductions of the different but physiologic effects of HCQ remain an appropriate average representation of HCQ’s effectiveness.

A second limitation is that we were unable to perform more robust analyses of the relationship between initiating HCQ treatment earlier and HCQ treatment effectiveness. In addition, the clinical settings of the trials varied, ranging from a focus on pre-exposure prophylaxis to early outpatient treatment. These differences may have affected our findings. However, they do represent a spectrum of outpatient COVID-19 transmission and illness, which is clinically relevant, and the variation in setting likely biased our meta-analysis toward the null.

## Conclusion

The randomized clinical trials performed to date demonstrate that hydroxychloroquine use in outpatients safely reduces the incidence of the composite of COVID-19 infection, hospitalization, and death.

### Patient and Public Involvement Statement

- At what stage in the research process were patients/public first involved in the research and how? NA: Patients and the public were not involved in this study because the study comprised an analysis of existing study data.
- How were the research question(s) and outcome measures developed and informed by their priorities, experience, and preferences? Various government, clinical and research entities have been trying to evaluate the degree of benefit of hydroxychloroquine in early COVID-19 outpatient treatment. Some individuals have stated publicly that results from randomized trials are needed for formal evaluation. We sought to identify and meta-analyze all relevant RCTs to date.
- How were patients/public involved in the design of this study? NA
- How were they involved in the recruitment to and conduct of the study? NA
- Were they asked to assess the burden of the intervention and time required to participate in the research? NA

## Data Availability

The original data extracted from the five studies analyzed herein are provided in the tables, figure and text. Readers are free to make use of these data.

https://doi.org/10.1101/2020.07.20.20157651

https://doi.org/10.1093/cid/ciaa1009

https://www.acpjournals.org/doi/10.7326/M20-4207

https://www.medrxiv.org/content/10.1101/2020.09.18.20197327v1

https://www.nejm.org/doi/full/10.1056/NEJMoa2016638

## Ethics Approval

Not required per se. The study comprised an analysis of existing study data, in which each of the original studies had already obtained institutional review board approvals.

## Transparency Statement

The lead author (the manuscript’s guarantor, HAR) affirms that the manuscript is an honest, accurate, and transparent account of the study being reported; that no important aspects of the study have been omitted; and that any discrepancies from the study as originally planned have been explained.

## Roles of the Authors

Idea for the study: HAR, PAM; database searching and study data extraction: JAL, HAR; analysis of data: HAR; interpretation of analysis results: JAL, PAM, JEM, HAR; writing initial draft of manuscript: JAL, HAR; critical review, editing and revision: JAL, PAM, JEM, HAR; controlling guarantor responsibility: HAR. All of the authors read and approved the final version of the manuscript, tables and figure.

## Key Messages Box

### What is already known on this topic

- Various government, clinical and research entities have been trying to evaluate the degree of benefit of hydroxychloroquine in early COVID-19 outpatient treatment.
- Seven nonrandomized but controlled clinical trials to date have shown significant reductions in hospitalization and mortality with early ambulatory hydroxychloroquine use, but individual randomized outpatient trials have not shown statistical significance of benefit with these or other outcomes.

### What this study adds

- The five outpatient randomized controlled studies to date examining new infection, hospitalization or mortality together show statistically significant evidence of reduced risk, RR=0.76 (95% CI 0.59 to 0.97).
- No serious adverse cardiac events were reported in any of the studies.
- The combined literature of seven nonrandomized controlled trials and five randomized controlled trials provides substantial and statistically significant evidence of benefit for early use of hydroxychloroquine in COVID-19 outpatients.

